# Large datasets from Electronic Health Records predict seizures after ischemic strokes: A Machine Learning approach

**DOI:** 10.1101/2024.01.24.24301755

**Authors:** Alain Lekoubou, Justin Petucci, Temitope Femi Ajala, Avnish Katoch, Souvik Sen, Vasant Honavar

## Abstract

**Objective:** To develop an artificial intelligence, machine learning prediction model for estimating the risk of seizures 1 year and 5 years after ischemic stroke (IS) using a large dataset from Electronic Health Records.

**Background:** Seizures are frequent after ischemic strokes and are associated with increased mortality, poor functional outcomes, and lower quality of life. Separating patients at high risk of seizures from those at low risk of seizures is needed for treatment and clinical trial planning, but remains challenging. Machine learning (ML) is a potential approach to solve this paradigm.

**Design/Methods:** We identified patients (aged ≥18 years) with IS without a prior diagnosis of seizures from 2015 until inception (08/09/22) in the TriNetX Research Network, using the International Classification of Diseases, Tenth Revision (ICD-10) I63, excluding I63.6 (venous infarction). The outcome of interest was any ICD-10 diagnosis of seizures (G40/G41) at 1 year and 5 years following the index IS. We applied a conventional logistic regression and a Light Gradient Boosted Machine algorithm to predict the risk of seizures at 1 year and 5 years. The performance of the model was assessed using the area under the receiver operating characteristics (AUROC), the area under the precision-recall curve (AUPRC), F1 statistic, model accuracy, balanced accuracy, precision, and recall, with and without anti-seizure medication use in the models.

**Results:** Our study cohort included 430,254 IS patients. Seizures were present in 18,502 (4.3%) and (5.3%) patients within 1 and 5 years after IS, respectively. At 1-year, the AUROC, AUPRC, F1 statistic, accuracy, balanced-accuracy, precision, and recall were respectively 0.7854 (standard error: 0.0038), 0.2426 (0.0048), 0.2299 (0.0034), 0.8236 (0.001), 0.7226 (0.0049), 0.1415 (0.0021), and 0.6122, (0.0095). Corresponding metrics at 5 years were 0.7607 (0.0031), 0.247 (0.0064), 0.2441 (0.0032), 0.8125 (0.0013), 0.7001 (0.0045), 0.155 (0.002) and 0.5745 (0.0095).

**Conclusion:** Our findings suggest that ML models show good model performance for predicting seizures after IS.

## Introduction

Stroke units, intravenous thrombolysis, and mechanical thrombectomy have transformed the prognosis of ischemic stroke, the most frequent stroke type.^1–3^ With more patients surviving the acute phase of stroke, there is an increased need for human and financial resources for post-stroke complications management. Seizure is a frequent and arguably underestimated complication of stroke. Prospective population-based studies estimate that over 10% of stroke survivors will cumulatively develop seizures following the index event in the next ten years.^4^ Late-onset post-stroke seizure risk factors include a cortical location, stroke volume, the presence of hemorrhagic transformation, and early seizure (seizure developing within 1-2 weeks of the stroke).^5,6^ Models designed to identify these predictors used limited variables and often assumed the potential association with post-stroke seizures. Hence, several predicting variables may remain to be identified. Hence, at the moment, at the individual level, it remains unclear who will develop seizures and who will not develop seizures. A recent systematic review of available risk prediction models revealed that when appropriately developed, available models did not include heterogeneous and relatively young populations as in the US.^7^ Such models may be challenging to use in busy clinical practices for post-stroke seizure risk classification. Besides the impracticability of such models, it may be challenging to allocate resources based on estimated post-stroke seizure risks at national and local levels. Further, previous anti-seizure treatment trials have failed partly because of inadequate assessment of individual stroke patients’ risk of seizures. Selecting high-risk patients for post-stroke seizure development from Electronic Health Records (EHR) remains nearly insurmountable. Thus, it is critical to have an appropriate risk model for post-stroke seizures, using routinely collected clinical information that is easily implementable to EHR. In this project, we aimed to use machine learning to aid in predicting the risk of seizures after ischemic stroke using a large number of variables in a large population from the largest network of electronic health records in the United States.

## Methods

### Data source

We identified patients with ischemic stroke without a prior diagnosis of seizures from 2015 until inception (08/09/22) in the TriNetX Research Network. The database collects de-identified information such as demographics, vital signs, diagnoses, procedures, patient location, medications, and mortality. In September 2022, 71 healthcare organizations (HCOs) participated in the database, with 106 million patients.

### Study population

We identified patients with stroke, aged ≥18 years, using the *International Classification of Diseases, tenth Revision* (*ICD-10*), from January 1, 2015, through August 9, 2022. Ischemic stroke patients were identified using ICD-10 I63 in EHR (excluding I63.6, which represents patients with cerebral infarctions due to venous thrombosis). All participants with a diagnosis of seizures at baseline were excluded. In all, we identified 430,254 participants with a diagnosis of stroke who met the study inclusion criteria.

### Assessment of outcome

The time at risk was 1-year and 5-year after the index stroke event. Seizures were identified using any of the ICD-10 codes G40 and G41.

### Covariates

Demographic variables included age (continuous variable), sex assigned at birth (male vs. female), and race/ethnicity. Race and ethnicity were grouped into four categories: Non-Hispanic White (NHW), Non-Hispanic Black (NHB), Hispanic, and others. Clinical variables included the following: diagnosis or history of hypertension, diagnosis or history of diabetes mellitus, diagnosis or history of atrial fibrillation, history of smoking, history of alcohol use, and stroke severity. The National Institute of Health Stroke Scale is the most commonly used score for stroke severity assessment; however, as this variable is not available in TriNetX Research Network, we assessed stroke severity using a combination of factors and variables described in the appendix. Treatment-related variables included the use of intravenous thrombolysis, mechanical thrombectomy, antiplatelet drugs, and anti-seizure drugs.

Procedure codes and codes used to identify drugs are available in the appendix file. Localization of ischemic stroke was divided into anterior cerebral artery strokes, middle cerebral artery strokes, and posterior cerebral artery strokes. Patients with concomitant diagnoses of brain pathologies or surgical interventions associated with an increased risk of seizures were identified using ICD-10 codes. These pathologies/surgical interventions included traumatic brain injury, benign brain neoplasms, malignant brain neoplasms, unspecified brain neoplasms, severe intracranial infection, bacterial meningitis, encephalitis, and decompressive craniotomy. ICD-10 and CPT codes were used to identify the variables. A list of all ICD-10 codes used is available in the supplemental materials.

### Machine learning model and interpretation: Machine learning model and interpretation

To develop and evaluate a generalizable model for seizure prediction at 1 year and 5 years, we used 5-fold nested cross-validation (CV) with non-overlapping training set (for training the model) and validation set (for hyperparameter tuning) and test set (for model evaluation). Initially, the entire dataset is stratified into 5 disjoint subsets, or folds. Subsequently, each fold is iterated over, serving once as the test set (red) while the remaining folds comprise the training set (blue). Within this training set, an inner cross-validation is conducted, dividing it into 5 further folds.

This step is crucial for hyperparameter optimization, where each parameter combination is trained on 4 folds (gray) and validated on the remaining fold (green), with the process cycling through all 5 folds to determine the best-performing hyperparameters. These optimal parameters are then used to train a new model on the full training set of the outer loop. The model’s predictive performance is assessed on the outer test set, ensuring each data point is used for testing just once. After completing all 5 iterations, the performance metrics across all 5 outer test sets are aggregated to produce a comprehensive evaluation of the model’s generalization capability (Figure 1). The following classification model types were explored: logistic regression, decision tree, random forest, LightGBM, AdaBoost, support vector machine, k-nearest neighbors, discriminant analysis, and Gaussian naïve Bayes.^8–14^ Scikit-learn was used to train and evaluate all models,^15^ except LightGBM, where Microsoft’s LightGBM library was employed.^16^ The LightGBM model yielded the best-generalized performance results. The hyperparameter optimization of LightGBM consisted of a grid search (within the nested cross-validation) over the tree depth, learning rate, and ensemble size. To account for the class balance between patients who developed seizures and those who did not, we used a cost-sensitive learning approach. The objective/cost function is modified to yield a stronger penalty for incorrectly predicting the minority class, i.e., those who developed seizures (by an amount proportional to the imbalance) using LightGBM’s *’class_weight=’balanced’* option. We used LightGBM and a set of feature importance scores derived from trained LightGBM models, and Shapley values ^17^ to determine the features most important in predicting seizures. We used Shapley values and partial dependence plots (PDP) to investigate the relationship between predictors and seizures. PDPs show only the average effect of the input variable, hence neglecting the impact of feature interactions, which can be present with tree-based models such as LightGBM.

**Figure 1:**
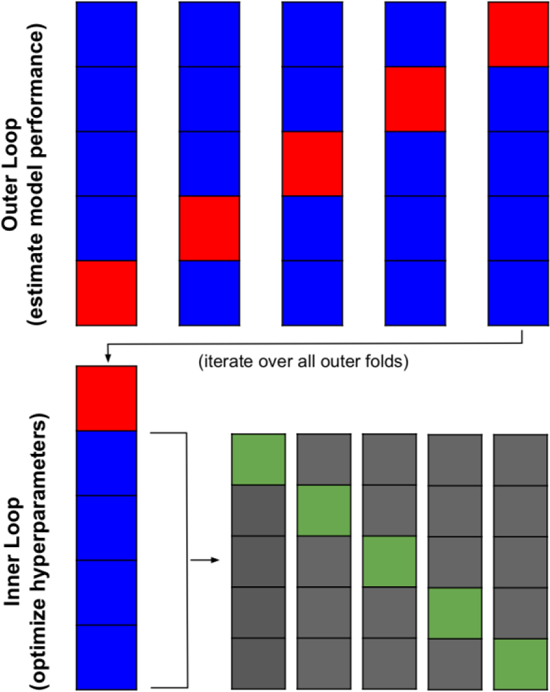
5-fold Nested Cross Validation. A visual representation of the nested 5-fold cross validation procedure. The outer loop partitions the data into 5 folds, where each fold serves as a unique and non-overlapping test set (red) once, while the remaining data forms the training set (blue). Within each outer training set, an inner 5-fold cross-validation is conducted, further dividing the data into 5 new folds. In this inner loop, one fold is used as the validation set (green) for hyperparameter tuning each iteration, and the other folds act as the training set (gray).

The performance of the model was based on the following metrics: area under the receiver operating characteristics (AUROC)-a chart that visualizes the tradeoff between true positive rate (TPR) and false positive rate (FPR), the area under the precision-recall curve (AUPRC-a curve that combines precision (PPV) and Recall (TPR) in a single visualization, the F1 statistic (it combines precision and recall into one metric by calculating the harmonic mean between those two), model accuracy (measures how many observations, both positive and negative, were correctly classified), balanced accuracy, precision, and recall. Performances were assessed, including excluding patients on anti-seizure drugs.

### Standard protocol approvals, registrations, and patient consent

This study protocol was submitted to the Pennsylvania State College of Medicine institutional review board and was not considered human subject research. All records contained within the database are fully de-identified. Thus, informed consent was waived.

## Results

General characteristics (Table 1 and Table 2): We included 430,254 patients with ischemic strokes and without a prior diagnosis of seizures. Seizures incidence at 1 year was 4.3% (18,502 patients) and 5.3% (22,675) at 5 years. The median age of patients who developed seizures was significantly lower compared to those who did not at one year and five years. The risk of seizures was similar between male and female stroke survivors. Patients who developed seizures were more likely to be Black individuals, smokers, have hypertension, large artery atherosclerosis disease, and develop severe stroke. Anti-seizure drug prescriptions, primarily Levetiracetam, were more frequent among those who developed seizures. They were less likely to have a posterior circulation stroke or receive intravenous tissue plasminogen activator or antiplatelet therapy. Patients who developed seizures were also more likely to be on electroencephalogram.

**Table 1:** Demographic and Clinical Characteristics.

| **Characteristics** | **Total Patient (%)**<br>**430,254** | **Seizure Incidence**<br>**following ischemic**<br>**stroke after 1 year**<br>**(N=18,502) 4.3%** | **Seizure Incidence**<br>**following ischemic**<br>**stroke after 5 years**<br>**(N=22,675) 5.27%** |
| --- | --- | --- | --- |
| **Age**, Median (SD), <sup>a</sup><br>years | 66.35± 14.9 | 64.2 ± 15.42 | 64.13 ±15.22 |
| **Sex** |  |  |  |
| Male | 225,871(52.5) | 9,766 (52.8) | 11,986 (52.9) |
| Female | 204,383 (49.1) | 8,736 (47.2) | 10,689 (47.1) |
| **Race** <sup>a</sup> |  |  |  |
| White | 274,882(63.9) | 10,925 (59.0) | 13,434 (59.2) |
| Black/ African<br>American | 73,914 (17.2) | 4,163 (22.5) | 5,173 (22.8) |
| Others or<br>Unknown | 81,458 (19.0) | 3,414 (18.5) | 4,068 (17.9) |
| **Stroke Risk** |  |  |  |
| Smoking <sup>a</sup> | 113,593 (26.4) | 5,793 (31.3) | 6,991 (30.8) |
| Hypertension <sup>a</sup> | 248,347 (57.7) | 12,054 (65.1) | 14,552 (64.2) |
| Diabetes <sup>a</sup> | 121,243(27.9) | 5,880 (31.8) | 7,084 (31.2) |
| Alcohol Use <sup>a</sup> | 20,156 (4.7) | 1,542 (8.3) | 1,821 (8.0) |
| Atrial fibrillation | 63,781(14.8) | 3,186 (17.2) | 3,746 (16.5) |
| Hyperlipidemia | 164,213 (38.2) | 7,468 (40.3) | 9,072 (40.0) |
| **Stroke Location** <sup>a</sup> |  |  |  |
| Anterior | 3,366 (0.8) | 205 (1.1) | 243 (1.1) |
| Posterior | 6,253 (1.5) | 358 (1.9) | 412 (1.8) |
| Middle | 32,863 (7.6) | 1,882 (10.2) | 2,267 (10.0) |
| ***Antiplatelet Therapy*** |  |  |  |
| Aspirin <sup>a</sup> | 170,384 (38.7) | 6,889 (37.2) | 8,374 (36.9) |
| Clopidogrel <sup>a</sup> | 60,300 (13.6) | 2,070 (11.2) | 2,565 (11.3) |
| Ticagrelor | 4,304 (1.0) | 164 (0.9) | 194 (0.9) |
| Prasugrel | 1,124 (0.3) | 39 (0.2) | 46 (0.2) |
| ***Electroencephalograms*** <sup>a</sup> |  |  |  |
| Continuous EEG 2-12 | 465 (0.1) | 117 (0.6) | 119 (0.5) |
| Continuous EEG 12-26 | 800 (0.2) | 207 (1.1) | 208(0.9) |
| Large artery Atherosclerotic Disease | 53,670 (12.5) | 3,053 (16.5) | 3,678 (16.2) |
| Intravenous TPA | 13,287 (3.1) | 651(3.5) | 775 (3.4) |

<sup>a</sup>These characteristics are significantly associated with seizure at both one-year and five years after a stroke. (*p-value* < 0.05)
TPA: Tissue Plasminogen Activator
EEG: Electroencephalogram
SD: Standard deviation

**Table 2:** Stroke severity and distribution of antiseizure medications.

| **Patient Characteristics** | **Total Number of patients**<br>**N=430,254 (%)** | **Seizure Incidence following ischemic stroke after 1 year**<br>**N=18,502 (4.3%)** | **Seizure Incidence following ischemic stroke after 5 years**<br>**N=22,675 (5.27%)** |
| --- | --- | --- | --- |
| ***Stroke severity*** |  |  |  |
| Aphasia | 39,596 (9.2) | 3,181 (17.2) | 3,645 (16.1) |
| Unspecific side hemiplegia | 5,232 (1.2) | 461(2.5) | 534 (2.4) |
| Non-dominant side hemiplegia | 21,835 (5.1) | 1,545 (8.4) | 1,821(8.0) |
| Aphasia | 31,711(7.4) | 2,857 (15.4) | 3,288(14.5) |
| Aphasia-Cerebrovascular | 5,137 (1.2) | 567 (3.1) | 661 (2.9) |
| Dysarthria | 28,291 (6.6) | 1,948 (10.5) | 2,259 (10.0) |
| Speech Disturbance | 23,786 (5.5) | 1,443 (7.8) | 1721 (7.6) |
| Facial weakness | 34,573 (8.0) | 2,252 (12.2) | 2,646 (11.7) |
| Cerebral<br>Intracranial edema | 643 (0.1) | 93 (0.5) | 113 (0.5) |
| Dominant left side<br>hemiplegia | 20,406 (4.7) | 1,408 (7.6) | 1,665 (7.3) |
| Altered mental<br>status | 46,840 (10.9) | 4,523 (24.4) | 5,074 (22.4) |
| Neurological<br>neglect syndrome | 5,181 (1.2) | 604 (3.3) | 672 (3.0) |
| Respiratory failure | 29,673 (6.9) | 3,201 (17.3) | 3,456(15.2) |
| Do Not Resuscitate | 16,893 (3.9) | 1,516 (8.2) | 1,577 (6.9) |
| Palliative care<br>consult | 11,904 (2.8) | 1,142 (6.2) | 1,169 (5.2) |
| Mechanical<br>thrombectomy | 4,164 (1.0) | 205 (1.1) | 253(1.1) |
| CE 1 | 25,383 (5.9) | 1,260 (6.8) | 1,557(7.0) |
| CE 2 | 39,282 (9.1) | 2,767 (15.0) | 3,222 (14.2) |
| Respiratory<br>ventilatory<br>procedure | 15,611 (3.6) | 2,067 (11.2) | 2,211(9.8) |
| Insertion of<br>endotracheal tube | 8,677 (2.0) | 1,168 (6.3) | 1,248 (5.5) |
| Insertion of feeding<br>device | 1,523 (0.4) | 162 (0.9) | 192 (0.8) |
| Intravenous<br>thrombosis | 13,287 (3.1) | 651 (3.5) | 775 (3.4) |
| ***Antiseizure drug <sup>a</sup>*** |  |  |  |
| Carbamazepine | 1,898 (0.4) | 419 (2.3) | 475 (2.1) |
| Clobenzepam | 157 (0.0) | 92 (0.5) | 97 (0.4) |
| Clonazepam | 9,317 (2.2) | 592 (3.2) | 722 (3.2) |
| Eslicarbazepine | 96 (0.0) | 76 (0.4) | 78 (0.3) |
| Ethosuximide | 67 (0.0) | 62 (0.3) | 64 (0.3) |
| Felbamate | 141 (0.0) | 64 (0.3) | 64 (0.3) |
| Gabapentin | 46,762 (10.9) | 2,202 (11.9) | 2,742 (12.1) |
| Lacosamide | 1,114 (0.3) | 504 (2.7) | 529 (2.3) |
| Lamotrigine | 3,298 (0.8) | 559 (3.0) | 634 (2.8) |
| Levetiracetam | 22,223 (5.2) | 5,910 (31.9) | 6,489 (28.9) |
| Oxcarbazepine | 1,385 (0.3) | 311(1.7) | 345 (1.5) |
| Perampanel | 88 (0.0) | 67 (0.4) | 70 (0.3) |
| Phenobarbital | 1,957 (0.5) | 308 (1.7) | 349 (1.5) |
| Phenytoin | 3,360 (0.8) | 1,002 (5.4) | 1,100(4.9) |
| Pregabalin | 8,834 (2.1) | 503 (2.7) | 615 (2.7) |
| Primidone | 1,368(0.3) | 140 (0.8) | 148 (0.7) |
| Rufinamide | 66 (0.0) | 62 (0.3) | 62 (0.3) |
| Tiagabine | 81 (0.0) | 62 (0.3) | 62 (0.3) |
| Topiramate | 5,721 (1.3) | 532 (2.9) | 640 (2.8) |
| Valproate | 4,608 (1.1) | 861 (4.7) | 951 (4.2) |
| Vigabatrin | 63(0.0) | 60 (0.3) | 60 (0.3) |
| Zonisamide | 541 (0.1) | 175 (0.9) | 189 (0.8) |

<sup>a</sup>These characteristics are significantly associated with seizure at both one-year and five years after a stroke. (*p-value* < 0.05)
CE 1(Clinical encounter 1): Describes a detailed interval history; A detailed examination; Medical decision making of high complexity.
CE 2 (Clinical encounter 2): Critical care, evaluation and management of the critically ill or critically injured patient; first 30-74 minutes

Figure 2 summarizes the performances of the model. Seven metrics are presented, including the area under the receiver operating characteristics (AUROC), the area under the precision-recall curve (AUPRC), the F1 statistic, model accuracy, balanced accuracy, precision, and recall, with and without seizure medication use in the models to allow an independent interpretation by the reader. These metrics were used simultaneously to account for the importance of classifying seizures and non-seizure patients and the heavily imbalanced sample. At 1 year, the AUROC, AUPRC, F1 statistic, accuracy, balanced accuracy, precision, and recall were respectively 0.7854 (standard error: 0.0038), 0.2426 (0.0048), 0.2299 (0.0034), 0.8236 (0.001), 0.7226 (0.0049), 0.1415 (0.0021), and 0.6122 (0.0095). Corresponding metrics at 5 years were 0.7607 (0.0031), 0.247 (0.0064), 0.2441 (0.0032), 0.8125 (0.0013), 0.7001 (0.0045), 0.155 (0.002) and 0.5745 (0.0095). The two most important features of the model were age and stroke in the middle cerebral artery territory (Figure 3). Partial dependent plots and cumulative probabilities of seizures for five selected features are presented in figure 4 and figure 5, showing an inverse relationship between age and the use of Aspirin but a positive relationship between seizures and the following variables: presence of altered mental status, middle cerebral artery location, and use of Levetiracetam.

**Figure 2:**
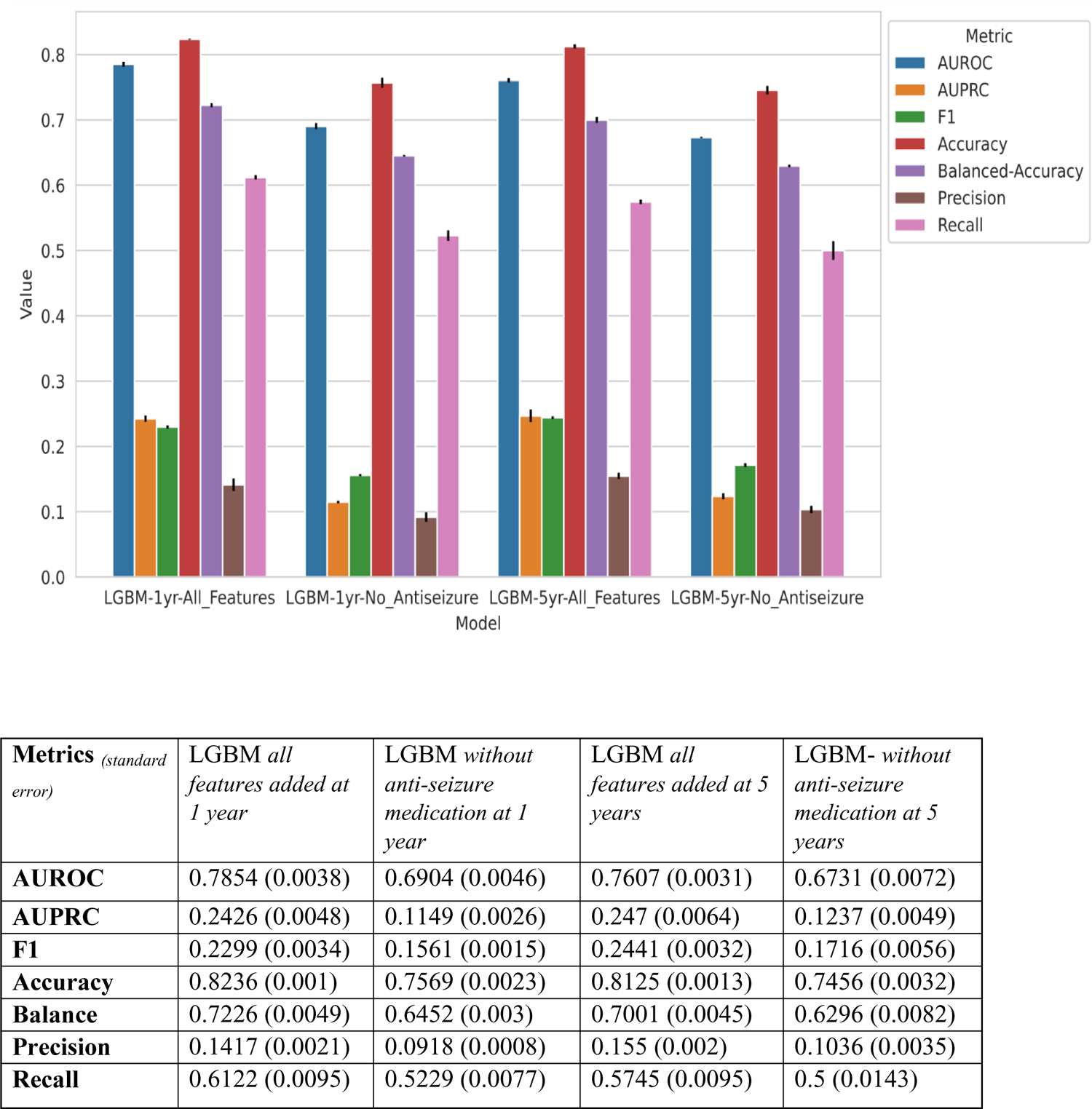
Visual Model Performance.

**Figure 3:**
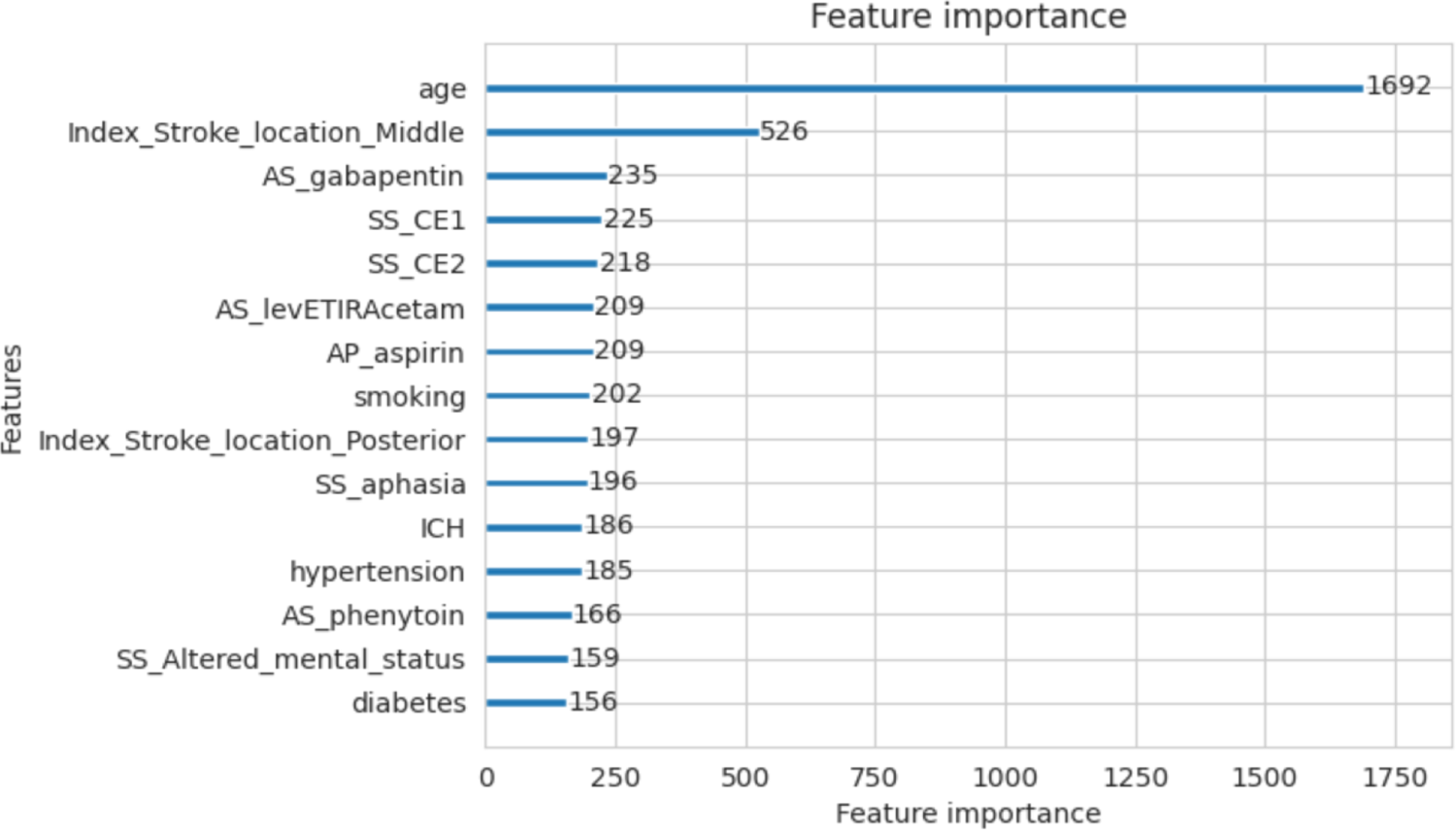
Top Important Features for LGBM Model. SS_CE1(Stroke Severity_Clinical Encounter 1): Describes a detailed interval history; A detailed examination; Medical decision making of high complexity. SS_CE2 (Stroke Severity_Clinical Encounter 2): Critical care, evaluation and management of the critically ill or critically injured patient; first 30-74 minutes. SS_aphasia: Stroke Severity_Aphasia ICH: Denotes patient with subsequent intracerebral hemorrhage ICD-10 codes. AS_: Ansitiseizure drugs.

**Figure 4:**
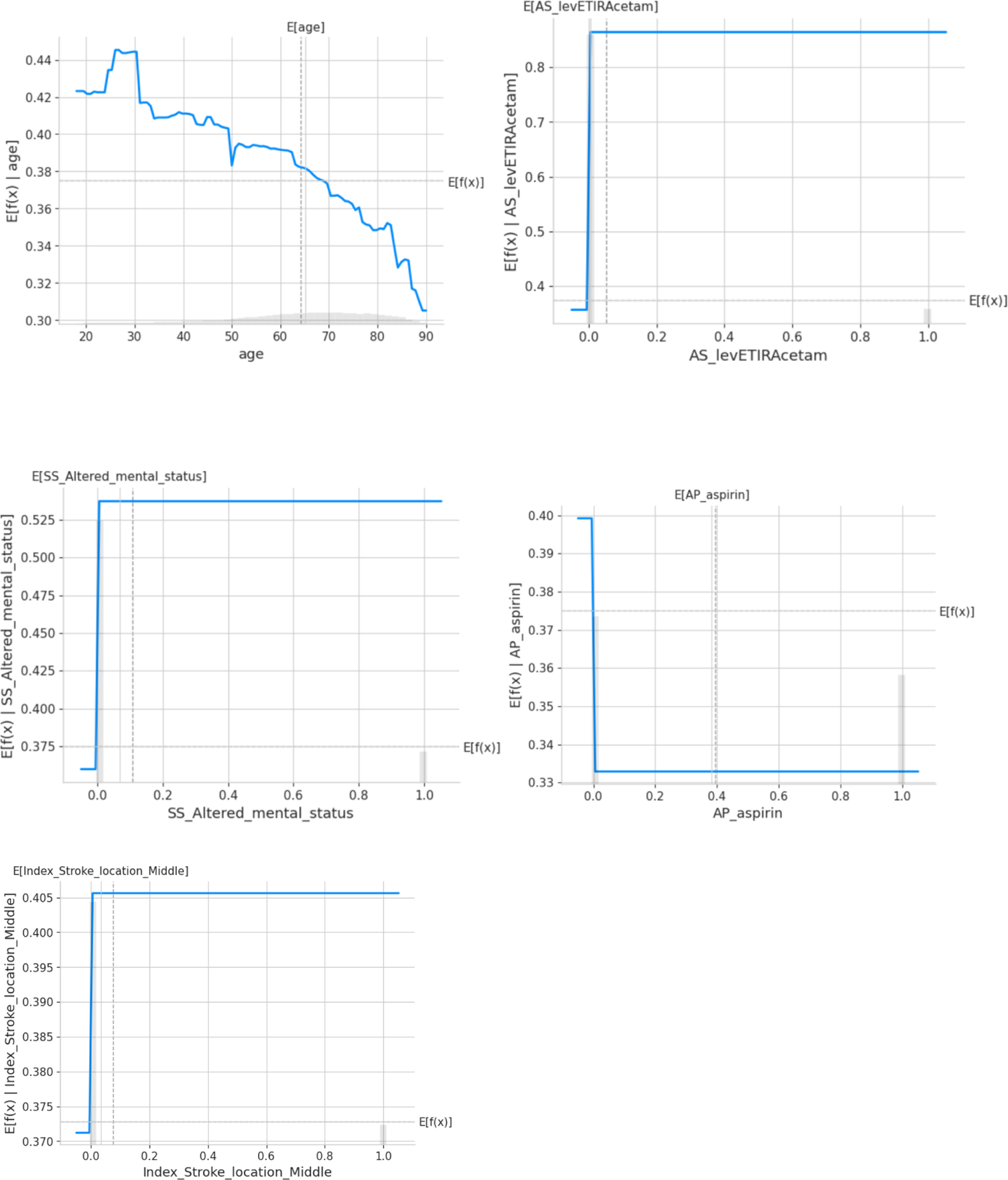
Partial dependence plots (PDP)

**Figure 5:**
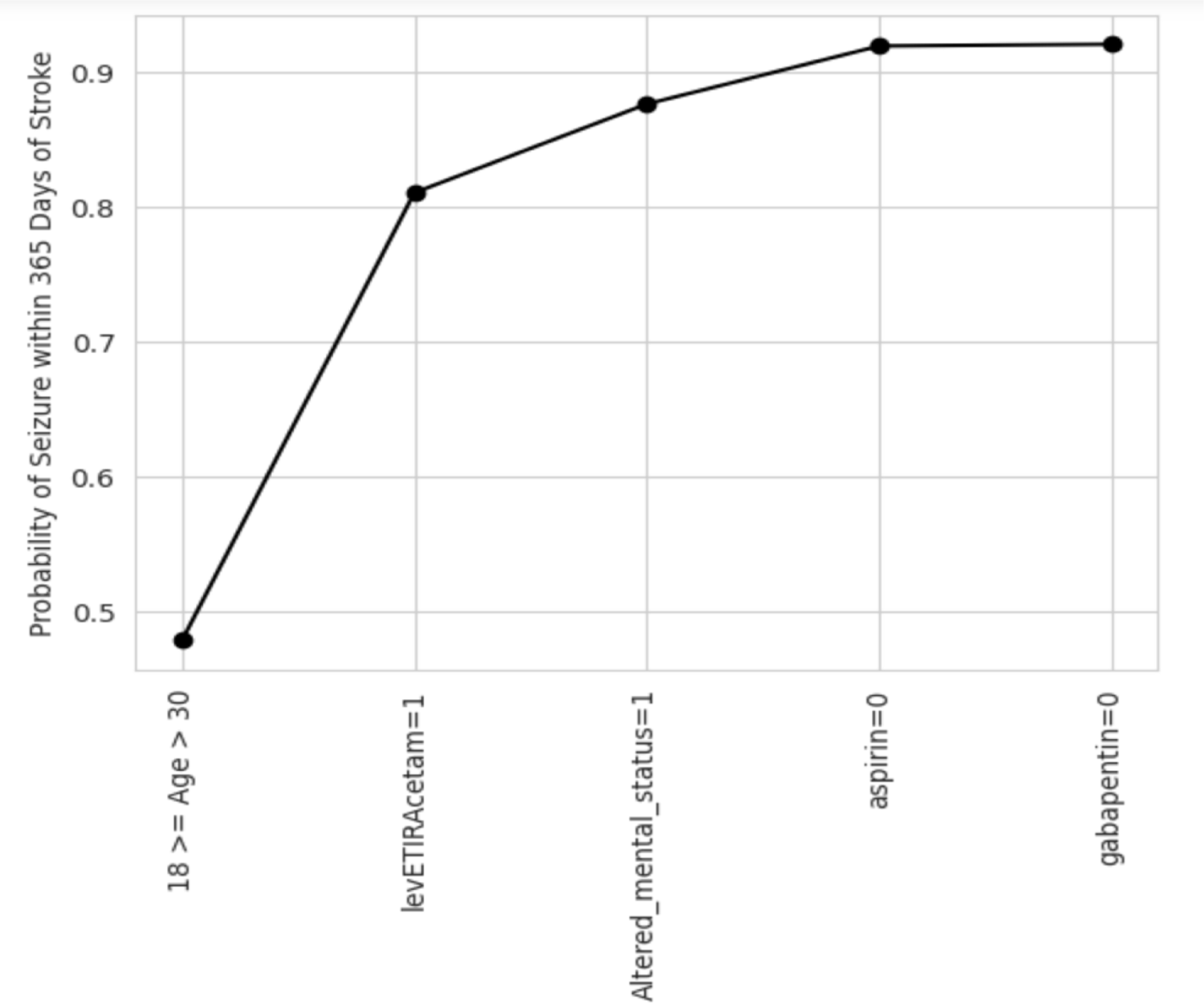
5 Feature Risk (Cumulative probabilities of Seizure Outcome) Model.

## Discussion

Our results indicate that the LightGBM model predicts seizures after ischemic stroke at 1 year and 5 years with a good performance using real-world data from a large sample of hospitalized patients across 71 healthcare organizations.

Seizures are frequent complications of ischemic stroke, yet strategies to prevent this complication are still to be developed and implemented. Preventing seizures is hampered by the lack of a generalizable predictive tool. Our systematic review conducted a year ago concluded that the only rigorously developed model was the SeLECT mode, a European-based model that predicted seizures with an area under the receiver operating characteristics of 77%. This model used the following granular clinical and imaging variables: the National Institute of Health Stroke Scale, the presence of early seizures, large vessel occlusion, cortical involvement, and involvement of the middle cerebral artery. Our model yielded similar performance as our AUROC at 1 year and 5 years were, 79% and 76%, respectively. We did not have access to granular data but used claims data, suggesting that seizure prediction is possible using data routinely collected in clinical practice. The model can be more easily integrated into electronic medical records, offering providers and researchers nearly instantaneous information on the risk of seizures after ischemic stroke. Further, our model was developed using data from a diverse US population, suggesting that it could be generalized to this population.

In this analysis, several features were associated with an increased risk of seizures in the LightGBM model. Age and strokes in the middle cerebral artery were the two most important features predicting seizures after ischemic stroke at 1 year and 5 years. The risk of seizures decreased with age, and strokes in the middle cerebral artery were associated with an increased risk of seizures. Counterintuitively, the findings of an increased risk of seizures in young individuals are in line with recent large-scale epidemiological data. For example, in two large datasets, including hospitalized patients in California, Florida, and New York, Stroke was more strongly associated with a subsequent seizure among patients <65 years of age compared to older patients.^18^ Similarly, in the population-based South London Stroke Register, younger patients were more likely to develop seizures than their older (65 years and above) counterparts.^19^ Stroke in the middle cerebral artery has also been reported as an important predictor of seizures after ischemic strokes.^20–22^

Our study has several implications. First, machine learning is a powerful tool that can be applied to large real-world datasets for predicting clinical outcomes such as seizures after stroke with accuracy similar to models developed using granular clinical data. It can, therefore, be purported that extracting and incorporating those granular data into Machine Learning-based models could lead to improved performances of those models. For example, natural processing language could be applied to electronic health records to extract such granular data, which in turn would improve the performance of a Machine Learning model, obviating the need to manually search Electronic Health Records, which would be impractical in large dataset such as the TriNetX Research Network. Second, we included 430,254 patients with ischemic strokes and without a prior diagnosis of seizures. This large number of patients and the use of Machine learning is more likely to detect predictors that would otherwise been missed using other approaches such as logistic regression analyses. This could explain why our model performed similarly to the best available model despite relying solely on Claim-based diagnoses and procedures. Third, our study has confirmed that a large dataset could resolve uncertainty about the association of some predictors with clinical outcomes. For example, in our study, age was the most important feature of our model, which was inconsistently associated with seizures in other studies of post-ischemic stroke seizures.^23–26^ Fourth, we observed that our models performed slightly less well when anti-seizure medications were not included as predictors. This highlights the complexity of using real-world data for model prediction and how different clinical scenarios can influence human factors (healthcare professionals) behaviors. For example, physicians may be more prone to prescribe anti-seizure medications to a patient who has epileptiform activity on the EEG or a patient with a large stroke. This has implications for future model development. It can be argued that accounting for patterns of human behavior will likely improve models’ performance and utility.

## Limitations

Our study has limitations that should be accounted for when interpreting its results. First, we relied on administrative ICD-10 diagnoses and procedures; therefore, we could not verify the accuracy of reporting these diagnoses. Second, stroke severity was not assessed using standard severity scales such as the National Institute of Health Stroke Scale; nevertheless, all proxy of stroke severity used in the current analysis were associated with an increased risk of seizures, suggesting the validity of our approach. Third, we did not have access to original imaging and EEG recording, which could have yielded additional predictors and improved the model’s performance. Despite these limitations, the use of a large sample across 71 healthcare organizations suggests that our results are likely to be generalizable to patients with ischemic stroke.

## Conclusion

In the current study, machine learning and, more specifically, LightGBM for Python library with sklearn API applied to the TriNetX Research Network, a large network of healthcare organizations in the United States, correctly predicted seizures after ischemic stroke at 1 year and 5 years about 75% of the time. The study shows the potential for applying machine learning to real-world data to predict seizures after ischemic strokes. Future studies will aim to improve the performances of ML-based models by incorporating more granular data, which could be identified from electronic health records using other features of artificial intelligence, such as natural language processing.

## Disclosure

None

## Data Availability

We used data from The TriNetX Research Network: HCOs (de-identified claims data), 106 million patients, which are available to researchers from participating centers.

